# Transcranial direct current stimulation (tDCS) targeting the medial prefrontal cortex (mPFC) modulates functional connectivity and enhances inhibitory safety learning in obsessive-compulsive disorder (OCD)

**DOI:** 10.1101/2021.02.16.21251796

**Authors:** Thomas G. Adams, Josh M. Cisler, Benjamin Kelmendi, Jamilah R. George, Stephen A. Kichuk, Christopher L. Averill, Alan Anticevic, Chadi G. Abdallah, Christopher Pittenger

## Abstract

**Background:** Psychotherapy based on fear extinction is a mainstay of treatment for obsessive-compulsive disorder (OCD). The default mode network (DMN) is important to safety signal processing, fear extinction, and exposure-based therapies. The medial prefrontal cortex (mPFC) is an anchor of the DMN. Neuromodulation targeting the mPFC might augment therapeutic learning and thereby enhance response to exposure-based therapies.

**Methods:** To characterize the effects of mPFC neuromodulation, 17 community volunteers completed resting-state fMRI scans before and after receiving 20 minutes of frontopolar multifocal transcranial direct current stimulation (tDCS). To examine the effects of tDCS on therapeutic learning, 24 patients with OCD were randomly assigned (double-blind, 50:50) to receive active or sham tDCS immediately before completing a two-day exposure and response prevention (ERP) challenge.

**Results:** After tDCS, frontal pole functional connectivity with regions in the anterior insula and basal ganglia decreased, while connectivity in the middle and superior frontal gyri increased (*ps*<.001, corrected). Functional connectivity between DMN and salience network (SN) increased after tDCS (*ps*<.001). OCD patients who received active tDCS exhibited more rapid within- and between-trial therapeutic extinction learning (*ps*<.05) during the ERP challenge compared to those who received sham tDCS.

**Conclusion:** tDCS targeting the mPFC may modulate SN and DMN functional connectivity and can accelerate therapeutic learning. Though limited by small samples, these promising findings motivate further exploration of the effects of tDCS on neural and behavioral targets associated with exposure-based treatments for OCD and for other anxiety and related disorders.

## Background

Approximately 28% of the American population will meet criteria for a disorder characterized by pathological anxiety during their lifetime.^1^ These disorders are often severe, intractable, and disabling.^2, 3^ Obsessive-compulsive disorder (OCD) afflicts approximately 1 in 40 Americans and has been ranked as a top 5 cause of disability among all mental illnesses.^4, 5^ Exposure-based cognitive-behavioral therapies are among the most efficacious treatments for OCD and other disorders of dysregulated anxiety.^6, 7^ However, partial response and relapse are common, and a sizeable minority of patients is treatment refractory.^8, 9^ Synergy between brain-based and psychotherapeutic strategies is an exciting avenue for the development of new therapeutic strategies to address this profound clinical need.^10, 11^

Exposure therapies depend on fear extinction.^12^ Extinction is a basic learning process whereby a fear is inhibited through the acquisition, consolidation, and recall of new “safety” learning.^13, 14^ Animal, human, and clinical research suggests that extinction learning occurs primarily within a circuit consisting of the medial prefrontal cortex (mPFC),^15^ hippocampus, and amygdala.^16-18^ Individuals with anxiety and related disorders, including OCD, have extinction learning deficits and abnormal recruitment of fear extinction circuitry, particularly the mPFC.^16, 19-25^ Moreover, deficient extinction learning, mPFC hypoactivity, and mPFC hypoconnectivity are associated with an attenuated response to exposure-based CBT.^26-34^

Recent work suggests that the default mode network (DMN), a network of functionally interconnected regions anchored by the mPFC, posterior cingulate cortex (PCC), and inferior parietal lobes,^35, 36^ supports the detection of safety cues and the acquisition and expression of safety learning.^37^ DMN connectivity is abnormal in a range of anxiety and related disorders, including OCD, and these abnormalities are associated with treatment response,^38-46^ including response to exposure-based CBT.^47-49^

These findings suggest that modulation of activity and/or long-term potentiation (LTP)-like plasticity within the DMN, particularly within the mPFC, may influence the processing of danger and safety cues, augment fear extinction, and ultimately improve the efficacy and efficiency of exposure-based CBT.^10, 14, 50^ Multiple groups have begun testing whether non-invasive brain stimulation of the mPFC can augment extinction learning and the efficacy of exposure-based therapies.^55,63,64^

Transcranial direct current stimulation (tDCS) is a safe, relatively inexpensive, and easily administered technique that can modulate the probability of neuronal firing by depolarizing or hyperpolarizing neurons under the anode and cathode, respectively.^51^ These effects can last up to 90 minutes after stimulation,^52-55^ due in part to the effects of tDCS on neurotrophic factors and LTP-like plasticity.^56^ tDCS has been shown to modulate functional connectivity of the area(s) being stimulated and augment functional connections between intrinsic networks.^57, 58^

Several previous studies have investigated whether tDCS can modulate extinction learning in humans, four of which indirectly targeted the vmPFC.^59-65^ Anodal tDCS applied before and during extinction training modestly enhances extinction acquisition,^59, 63, 65^ while anodal tDCS applied during the consolidation of extinction learning (after training) marginally improved extinction recall after a 24-hour delay.^60^ One study applied anodal tDCS over the frontal pole (anode over nasion/Fpz) *during* extinction training and found no beneficial effects on the acquisition or recall of extinction learning, compared to sham tDCS.^64^ The ability of tDCS to enhance extinction learning via mPFC modulation therefore remains unclear.

Another limitation of this literature is that it investigates the extinction of fear conditioning administered in the laboratory, which may not engage the same mechanisms as chronically dysregulated anxiety in neuropsychiatric disorders. A single study has extended this work to extinction of neuropsychiatric symptomatology. van ‘t Wout and colleagues recently reported the effects of tDCS targeting the vmPFC (via the ventrolateral PFC) on therapeutic learning during virtual reality exposure among 12 military veterans with PTSD.^66^ Compared to sham tDCS, veterans who received anodal tDCS (2mA) simultaneously with six sessions of VR exposure evinced accelerated between-trial reductions in physiological arousal and reduced PTSD symptomatology.^66^

No studies of tDCS in extinction learning have reported on the neural effects of the brain stimulation protocol used, and all have used bipolar tDCS: a single anode and a single cathode. In bipolar tDCS, electrical flux is greatest beneath the anode and cathode, but the passed current also has significant effects on most of the surrounding and intervening neural tissue.^67, 68^ Indeed, this is what has allowed previous work to indirectly target the vmPFC, via electrodes placed on the ventrolateral PFC and contralateral mastoid process. Multifocal or high-definition tDCS provides far greater spatial precision than bipolar tDCS by using more than two electrodes to deliver stimulation, typically one stimulation electrode and four or more return electrodes.^67, 68^ Modeling of electrical fields produced by multifocal tDCS suggest that the effects of the stimulation electrode are concentrated below the electrode, while the effects of the return electrodes are small and distributed.^68^ This comports with neuroimaging data.^68^

We tested the effects of multifocal tDCS targeting the mPFC on resting functional connectivity and on extinction learning during therapeutic exposure in patients with OCD. In Study 1, community volunteers completed resting-state functional magnetic resonance imaging (rs-fMRI) scans before and after administration of frontopolar multifocal tDCS. In Study 2, OCD patients received active or sham frontopolar multifocal tDCS immediately before completing 50 minutes of individualized *in vivo* exposure exercises to examine the effects of tDCS on therapeutic extinction learning.

### Methods: Study 1

Eighteen adult community volunteers were recruited from the greater New Haven area. Volunteers completed a brief phone screen to probe for common psychiatric diagnoses; formal diagnostic interviews were not conducted. After providing informed consent and completing a Yale Human Investigations Committee (HIC/IRB)-approved informed consent form, volunteers completed standard tDCS and MRI safety screening tools,^69, 70^ a demographic form, and self-report ratings of depression, anxiety, and OCD (**Supplementary Table 1**).^71, 72^ Participants then underwent structural MRI and baseline resting state fMRI (rs-fMRI) scans (details below). Volunteers were then escorted to a nearby room to receive 20-minutes of tDCS targeting the mPFC (details below). Immediately following tDCS, volunteers were returned to the scanner bay to complete a post-tDCS rs-fMRI scan. Participants completed the *tDCS Adverse Effects Questionnaire*,^70^ and compensated $50 for their time.

**Table 1.**
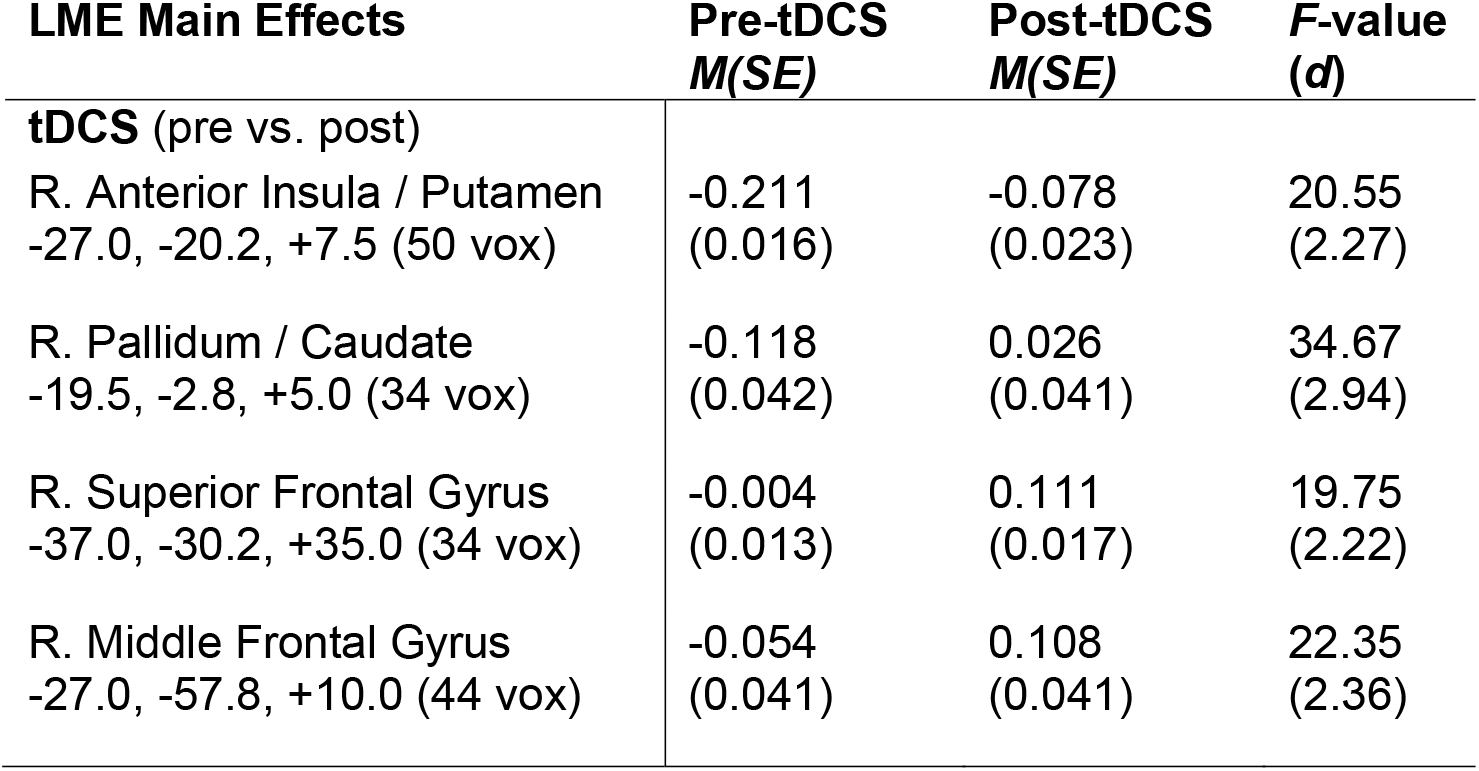
Summary of peak areas of change in functional connectivity with the frontopolar seed from pre- to post-tDCS (cluster corrected [> 30.6 voxels], *p* < .001). Cluster size (# voxels) and x,y,z coordinates (MNI) provided.

#### MRI Acquisition and Processing

Imaging data were collected on a Siemens Prisma 3T system using a 64-channel foam-padded phased array head coil. An HCP-style multiband fMRI sequence was used for rs-fMRI acquisition. Two runs of approximately six minutes (504 volumes) were completed before and after tDCS. Participants were instructed to rest with their eyes open during scanning and were video monitored to ensure they stay awake. Image preprocessing was completed using AFNI software using standard steps as previously reported.^73^ See Supplemental Materials for sequence and processing details.

#### Transcranial Direct Current Stimulation (tDCS)

tDCS was delivered using a battery driven Starstim transcranial electric stimulator (Neuroelectrics®, Cambridge, MA) through 1cm^2^ ceramic electrodes. Twenty minutes of facilitatory multifocal tDCS targeting the mPFC was administered to all volunteers. Current was ramped in and out for 30 seconds at the beginning and end of stimulation to mitigate sensory effects. A single anode was placed over the frontal pole (Fpz [10-20 EEG] at 1.5mA) and five return electrodes (cathodes) were arranged in a circumferential array (AF7, AF8, F3, F4, and Fz at 0.3mA, **Figure 1a**). Modelling of electrical field distribution suggests that this electrode montage concentrates positive current in the frontal pole beneath the anode, while negative current is of limited strength beneath the five cathodes (**Figure 1b**).

**Figure 1.**
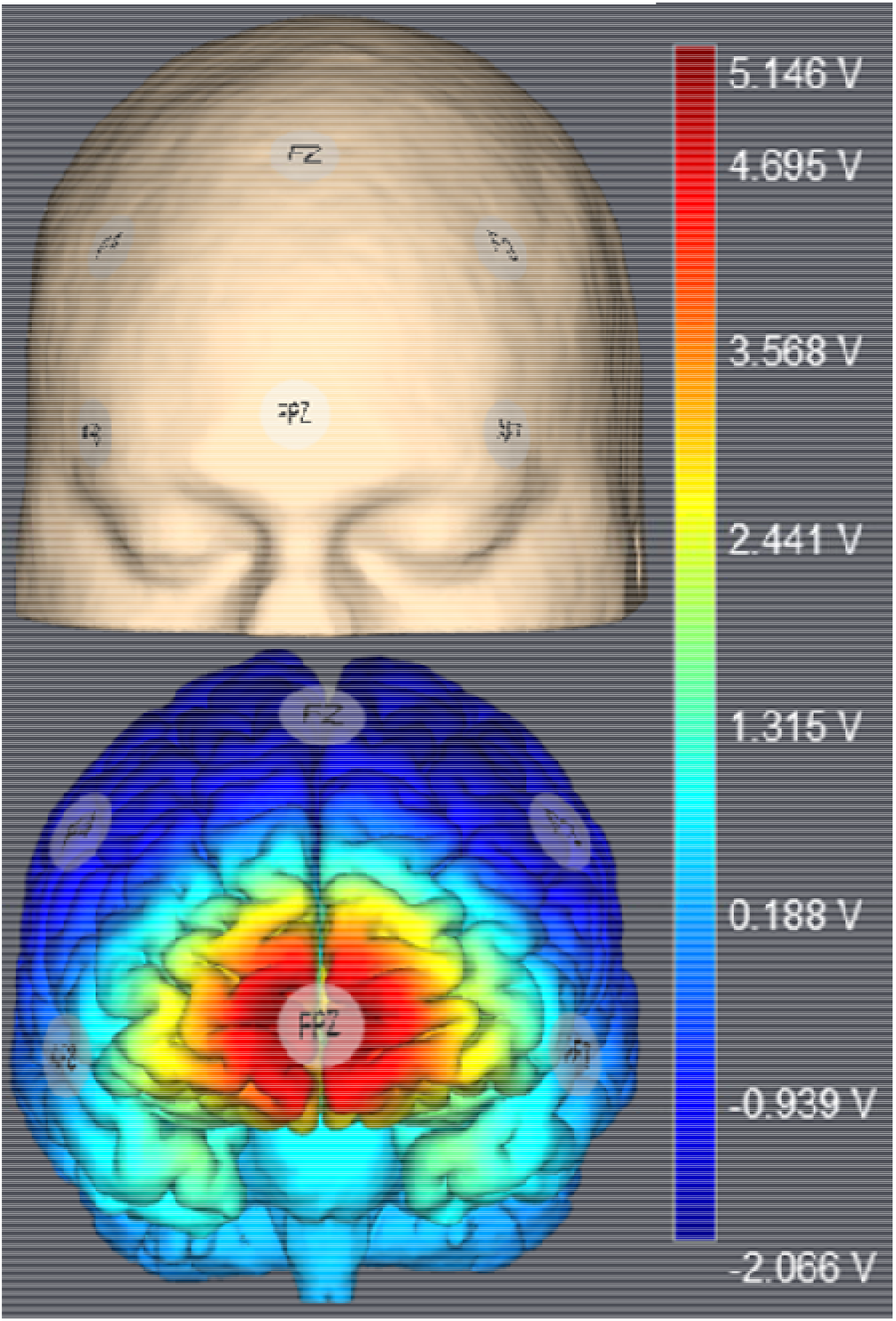
1.5 mA multifocal tDCS was administered using a Starstim transcranial electric stimulator, with a 1 cm^2^ anode over Fpz (10-20 EEG) surrounded by five cathodes in a circumferential array (AF3, AF4, F3, FZ, and F4). Simulation of the electrical fields produced by this montage was performed using Stimweaver and showed enhance electrical field potentials throughout the mPFC, particularly the frontopolar cortex and adjacent cortices, with limited effects on surrounding grey matter, including in brain tissue beneath the cathodes.

### Methods: Study 2

Twenty-six individuals with a primary diagnosis of OCD were recruited from the greater New Haven community for Study 2. All interested individuals first completed a phone screen to determine preliminary eligibility. After passing the initial phone screen, volunteers were invited to complete an in-person screening intake that included a psychiatric interview with or supervised by a doctoral-level clinician. Evaluations includes, a structured diagnostic interview [Mini International Neuropsychiatric Interview (MINI)^74^ or Structured Clinical Interview for DSM-IV Axis I Disorders (SCID-I)^75^], and the Yale-Brown Obsessive-Compulsive Scale (YBOCS) Checklist and Severity Scale.^76, 77^ Co-morbid psychosis, autism, mania, active substance abuse, and major neurological disease were grounds for exclusion. Participants were medication free or stably (≥ 4 weeks) medicated with an SSRI. Anxiolytics (particularly benzodiazepines) can interfere with extinction learning and exposure therapy and thus were grounds for exclusion,^78, 79^ as were medications that might lower seizure threshold (e.g., psychostimulants).^80^

On the first day of the experiment, OCD patients provided informed consent and signed a Yale HIC/IRB-approved consent form. They then completed standard tDCS safety screening tools,^69, 70^ a demographics form, and self-report ratings of depression, anxiety, and OCD (**Supplementary Table 1**).^71, 72, 81^ To complete the *ERP Challenge* OCD patients were provided standard ERP psychoeducation and were taught to use a 0-to-100-point subjective units of distress scale (SUDS) to rate their emotional distress throughout the experiment. Next, OCD patients began the two-day *ERP Challenge*, which was used to assess therapeutic extinction learning.^82^ On each day of the ERP Challenge, OCD patients completed five 10-min. trials of an individualized *in vivo* exposure exercise (see **Supplemental Table 2** for example exercises). SUDS was reported every minute of the ERP Challenge to monitor within- and between-trial therapeutic extinction learning. See Supplemental Materials for full details. OCD patients were randomized to receive Active-(as per Study 1) or Sham-tDCS (see below) before completing 50 minutes of individualized exposure. The patient and experimenter administering the ERP Challenge were blind to tDCS condition. Day 1 concluded with completion of the *tDCS Adverse Effects Questionnaire* ^*70*^ and a single 5-point scale to assess the degree to which they believed that they received real (active) or placebo (sham) tDCS. OCD patients returned 18-36 hours later to complete a second day of the same exposure exercises to examine the return of fear; no tDCS was administered on Day 2. After this second exposure exercise, participants were debriefed and compensated $80 for their time.

**Table 2.** Fixed effect estimates (linear mixed effects model) of the intercept and the main effect of tDCS on connectivity between the salience network (SN) and subnetworks of the default mode network (DMN) and a cingulo/SMA network. All effects are significant at *p* < .05 with Bonferroni correction for familywise error (*p*s ≤ .005).

| **(Inter-Network Connectivity)** | **Intercept**<br>***B* (*SE*)** | **tDCS**<br>***B* (*SE*)** | **Upper - Lower**<br>**C.I. (95%)** | **Cohen's *d*** |
| --- | --- | --- | --- | --- |
| SN to Anterior DMN | -0.21 (0.03) | 0.05 (0.02) | 0.02 to 0.09 | 0.73 |
| SN to Posterior DMN | -0.30 (0.03) | 0.05 (0.02) | 0.02 to 0.08 | 0.77 |
| SN to Ventral/Medial DMN | -0.12 (0.03) | 0.06 (0.02) | 0.03 to 0.09 | 0.94 |
| SN to Cingulo/SMA | 0.32 (-.03) | -0.05 (0.01) | -0.07 to -0.19 | 0.85 |

**Table 3.** Linear mixed effects modeling of within- (mins) and between-trial (trials) learning during Day 1 of exposure with response prevention (ERP) challenge among OCD patients who received active tDCS (*n* = 12) or sham tDCS (*n* = 12) immediately before completing five 10-min trials of *in vivo* exposure. The significant group*mins and group*trials interactions identify accelerated within- and between-trial extinction learning in the active tDCS group relative to the sham tDCS group (see **Figure 4**).

| Fixed Factors | Beta (SE) | 95% C.I. |  |
| --- | --- | --- | --- |
|  |  | Lower | Upper |
| Intercept | 51.41 (4.40) | 42.33 | 60.48 |
| Mins | -.49 (.31) | -1.12 | 0.14 |
| Trials | -2.47 (1.30) | -5.15 | 0.21 |
| Group | -5.54 (6.22) | -18.37 | 7.30 |
| Mins*Trials | -.01 (.06) | -0.12 | 0.11 |
| Group*Mins | -1.67 (.44)** | -2.56 | -0.78 |
| Group*Trials | -3.83 (1.84)* | -7.63 | -0.04 |
| Group*Mins*Trials | .31 (.08)** | 0.15 | 0.46 |

Note; \* = $p < .05$ ; \*\* = $p < .01$ . SUDS = Subjective Units of Distress

#### Transcranial Direct Current Stimulation (tDCS)

Active-tDCS procedures were identical to those described for Study 1. Sham-tDCS used the same multifocal montage, but current was ramped in/out for 30 seconds at the beginning and end of a 20-minute period during which no current was delivered. Both the OCD patients and the experimenter administering the ERP Challenge were blind to experimental assignment, though the staff administering the tDCS was not.

### Results: Study 1

*Analytic Strategy:* Functional connectivity to a seed region beneath the point of anodal stimulation was computed following previously published procedures and standard AFNI software (see **Supplemental Materials**).^83^ A linear mixed effects model (LME) was calculated using 3dLME to estimate the effects of tDCS on each voxel’s connectivity with this seed region. The main effects of tDCS (pre vs. post) and scan run (first vs. second) and the tDCS*Run interaction were modelled as fixed effects and the intercept was modelled as a random effect.

Cluster-level thresholding (3dclustersim) using an autocorrelated function (-acf in 3dFWHMx) was applied to the resulting LME group-level effects to control for voxelwise comparisons with a corrected *p* < .05 (two-tailed given an uncorrected *p* < .001).^84^ This resulted in a necessary cluster size of 30.6 face-connected voxels (NN1). Cluster size, coordinates, descriptives, and peak *F*-values for all significant clusters are presented in **Table 1**.

There was a significant main effect of tDCS on functional connectivity within four clusters, all of which were in the right hemisphere, and all four of which shifted towards less negative/more positive functional connectivity following tDCS (**Figure 2A**). Negative functional connectivity between the frontal pole and two clusters centered in the basal ganglia and insula decreased following administration of tDCS (**Figure 2B**). A cluster comprising regions of anterior insula and putamen reduced in negative connectivity, while another cluster within the pallidum and caudate changed from negative to positive connectivity. Functional connectivity between the frontal pole and clusters centered in the right middle frontal gyrus (mFG) and superior frontal gyrus (sFG) increased following administration of tDCS (**Figure 2B**); both clusters changed from negative to positive connectivity.

**Figure 2.**
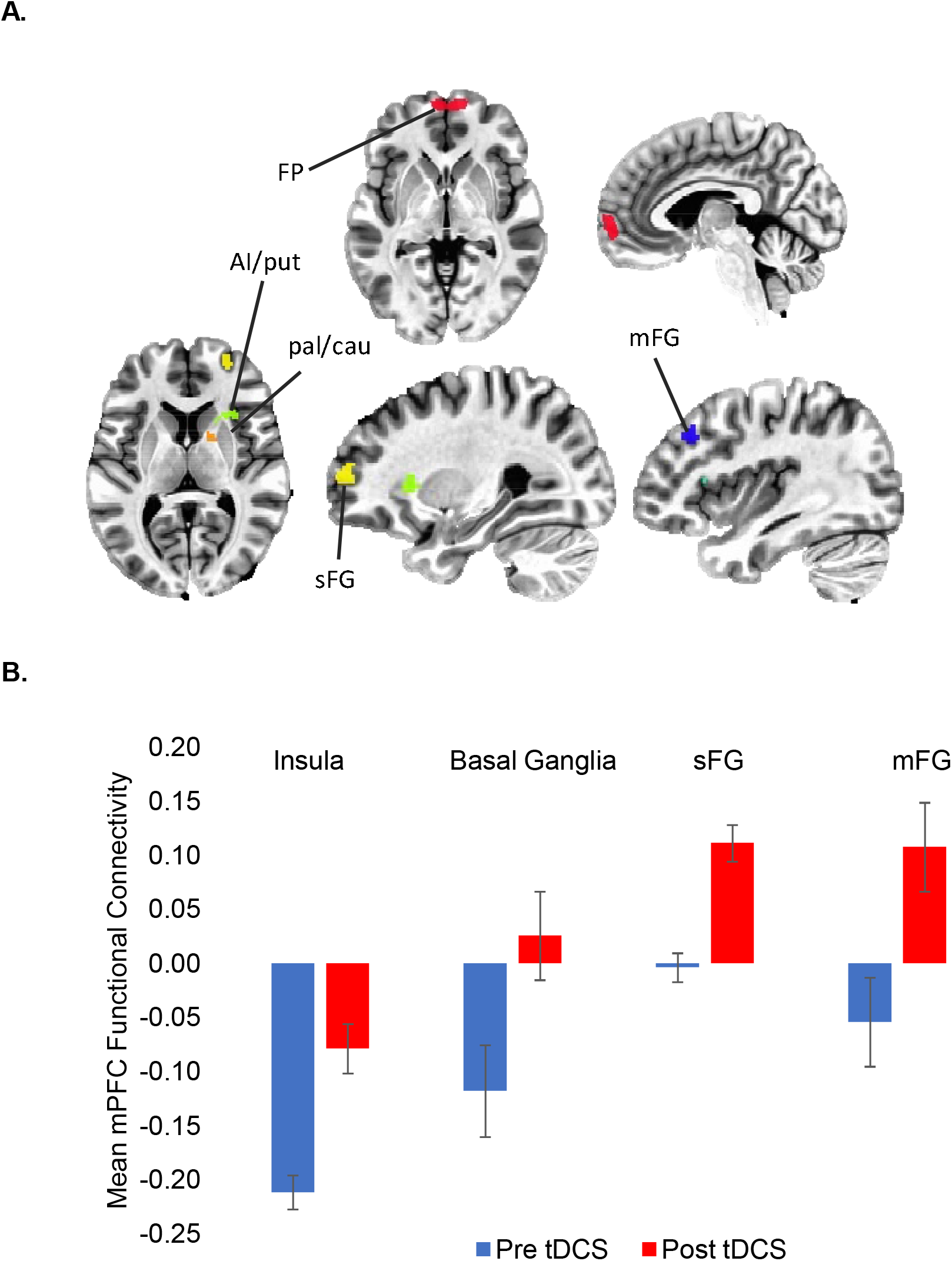
**(A)** Seed-based functional connectivity analyses with stringent cluster correction (*p* < .001, cluster sizes > 31 voxels) before and after administration of tDCS identified reduced negative functional connectivity between the frontopolar (FP) seed (red) and the right anterior insula / putamen (AI/put; green) and pallidum / caudate (pal/cau; orange), and increased functional connectivity between the FP and the right superior and middle frontal gryi ([sFG] blue and [mFG] yellow). (**B**) Mean connectivity between FP and each of these clusters before and after tDCS.

Inspection of significant subcortical clusters suggested functional connectivity was reduced between the frontal pole and reward circuitry, particularly the anterior striatum (**Supplemental Figure 2**). Inspection of significant cortical structures suggested that the frontopolar seed mapped exclusively onto the DMN, the frontal sFG and mFG clusters mapped primarily onto the executive control network (ECN), and the anterior insula cluster mapped primarily onto the salience network (SN; **Supplemental Figure 3**).

**Figure 3.**
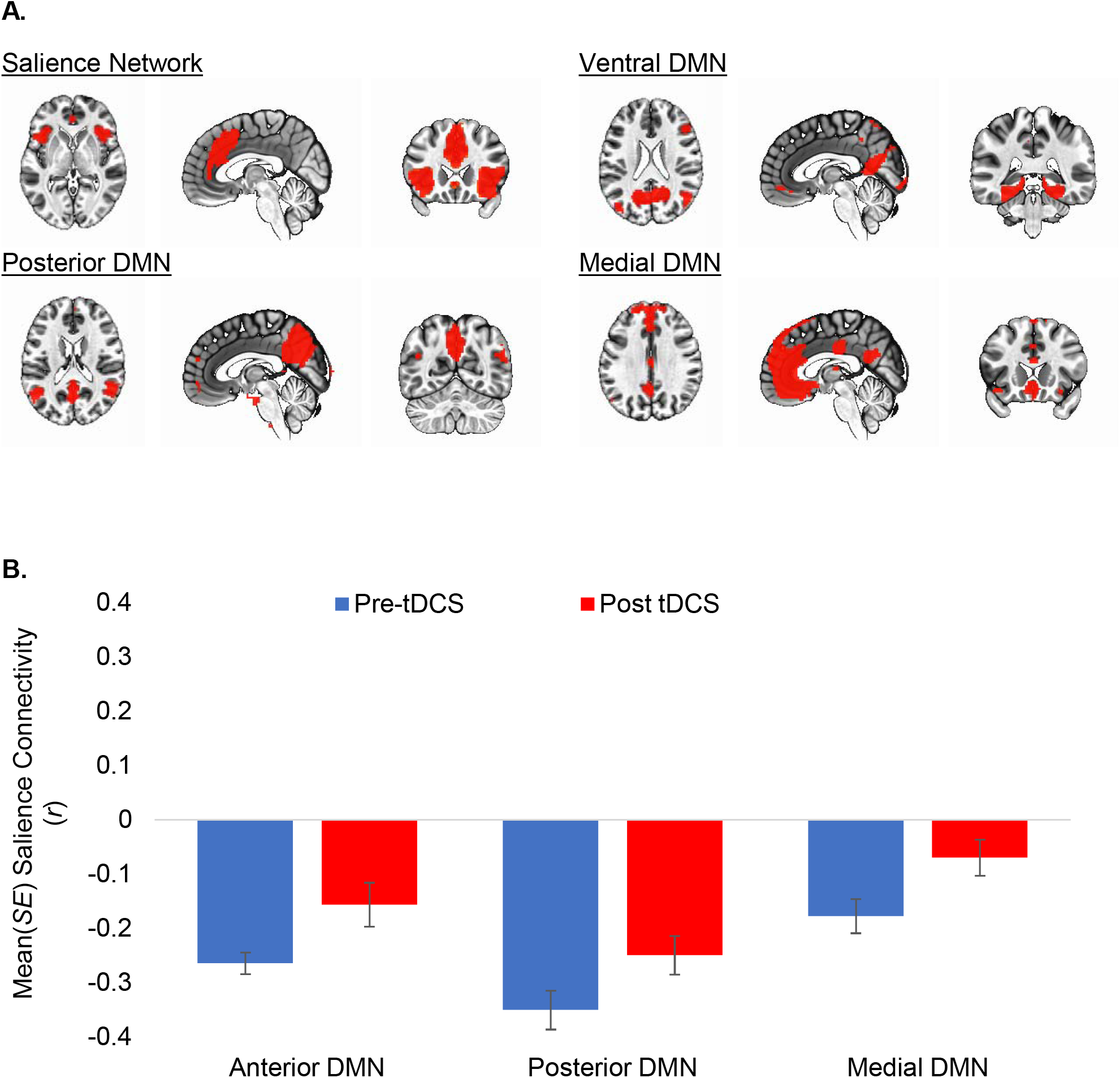
Inter-network connectivity between the salience network (SN) and subnetworks of the default mode network ([DMN] ventral, posterior, and medial) significantly (*ps* < .005, significant after Bonferroni correction) decreased following administration of frontopolar tDCS.

To test whether the effects of tDCS were truly a lateralized effect or is simply an artifact of thresholding, we conducted exploratory analyses with relaxed voxelwise correction (uncorrected *p* < .01). In addition to the previously listed clusters in the right hemisphere, functional connectivity changes were observed in 12 additional clusters, including between the frontal pole and the left superior and middle frontal gyri and the left basal ganglia. This suggests that most effects of tDCS are likely bilateral, though statistically stronger on the right. See **Supplemental Figure 1**.

To explore the effects of frontopolar anodal tDCS on inter-network connectivity, we conducted independent component analysis (ICA) of resting state data using previously published methods (see **Supplemental Materials**).^73, 85, 86^ Data were combined across all four scan runs for estimation of independent components to allow for within-subject (pre and post tDCS) contrasts of inter-network connectivity. To identify networks for second-level analyses, all functional networks were visually inspected for overlap with the mPFC or significant clusters from the seed-based analysis; artifact networks (e.g., motion or ventricle networks) were discarded. This resulted in the selection of seven networks: the SN, the right ECN (frontoparietal), three components of the DMN (ventral, posterior, medial), and two striatal networks. Linear mixed effects models (MATLAB fitlme.m) were used to examine the effects of tDCS on connectivity between the selected networks; intercept, tDCS, run, and tDCS*run were modelled as fixed effects and the intercept was modelled as a random effect. Bonferroni adjustment was used to correct for familywise error (*p* ≤ .007).

The main effect of tDCS was significant in three models, all of which included the SN and a DMN component (**Figure 3A**). Negative intercept coefficients indicate that the SN was anticorrelated with all three DMN components at baseline (pre-tDCS). Significant positive main effects of tDCS indicate that these anticorrelations were reduced following administration of tDCS (*ps* ≤ 0.005; **Table 2** and **Figure 3B**). The main effects of tDCS for all remaining contrasts were non-significant following correction for multiple comparisons (*p*s > 0.007), including contrasts between the SN and the right ECN and the two striatal networks.

### Results: Study 2

We next tested whether multifocal tDCS targeting the frontal pole can enhance therapeutic extinction learning in patients with OCD. Participants were randomized to receive either active (real) or sham (placebo) tDCS before the first of two one-hour individualized exposure challenges, which were modeled on standard therapeutic exercises during exposure and response prevention (ERP) therapy (see Methods).

All analyses were completed using SPSS 22.^87^ There were no significant differences in age or the distribution of gender and race between active and sham tDCS groups (all *ps* > .10; **Supplemental Table 1**). There were also no significant differences in the severity of OCD, anxiety, or depressive symptoms, nor were there differences in the percent of those on psychotropic medications at the time of the experiment (all *ps* > .10; **Supplemental Table 1**). There were no significant differences in SUDS ratings (*p*s > .10) immediately prior to or following administration of tDCS (but before beginning exposure; **Supplemental Figure 3A**). Importantly, both groups reported that they believed they were receiving Active stimulation, with no difference in confidence between groups (**Supplemental Figure 3B**), confirming that our Sham condition and blinding were effective.

Linear mixed effects (LME) modeling was used to examine the effects of tDCS on within- and between-trial therapeutic learning during Day 1 of the ERP Challenge (see **Table 2**). The intercept, mins (1-10), trials (1-5), group (sham tDCS [0] vs. active tDCS [1]) and all two-and three-way interactions were modeled as fixed effects. The intercept, mins, and trials were also modelled as random effects; only fixed effects are reported. There was a significant group*mins*trials interaction (*β* = 0.31, 95%C.I. = 0.15 to 0.46), suggesting that the mins*trials interaction varied by experimental group. Within-trial reductions in SUDS were greater for OCD patients in the active tDCS than in those in the sham-tDCS group (**Figure 4**; group*mins *β* = - 1.67, 95%C.I. = -2.56 to -0.78). Similarly, between-trial reductions in SUDS were greater for OCD patients in the active tDCS group than in the sham-tDCS group (**Figure 4**; group*trials *β* = -3.83, 95%C.I. = -7.63 to -0.05). On average, OCD patients in the active tDCS group reported a 64.6% reduction in SUDS from the beginning of the first trial to the end of the last trial of the ERP Challenge on Day 1, whereas those in the sham tDCS group reported only a 24.2% reduction (*t*[22]=5.58, *p* < .01).

**Figure 4.**
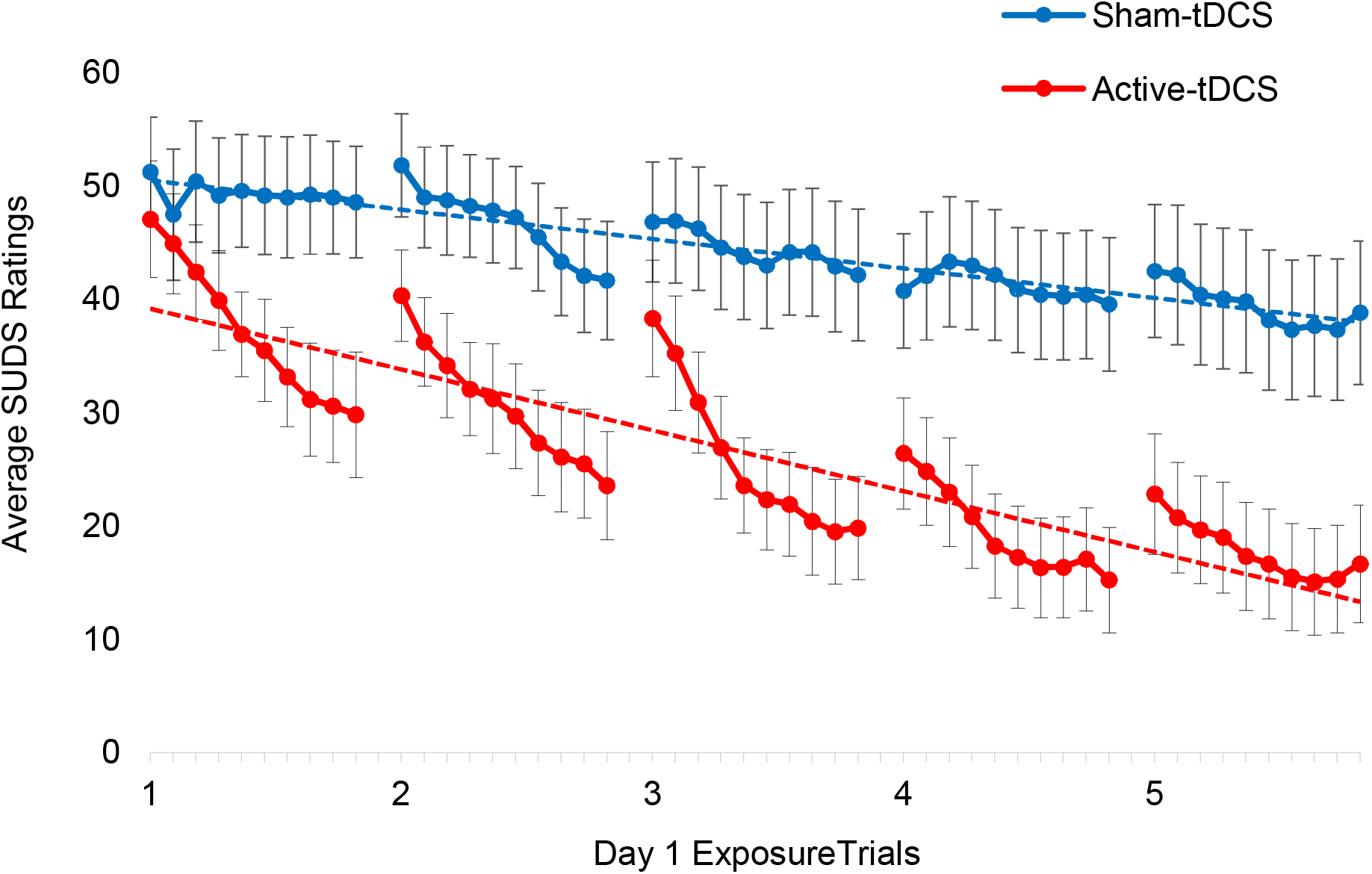
Despite nearly identical SUDS ratings at the beginning of the ERP Challenge (min. 1 of trial 1), OCD patients who received active tDCS reported significantly greater within-trial (solid lines, *p* ≤ .05) and between-trial (dashed lines, *p* ≤ .01) therapeutic extinction learning than those who received sham stimulation. ***Note***. SUDS = Subjective Units of Distress (0-100)

A LME model was used to examine the return of fear on day 2. The intercept, group, and time were all modelled as fixed effects; time was defined as SUDS ratings from the end of the last exposure trial on Day 1 (coded as 0) and the beginning of the first trial on Day 2 (coded as 1). Return of fear was similar across the two experimental groups (group*time *β* = 10.83, 95%C.I. = −8.24 to 29.91).

## Discussion

We found that multifocal anodal frontopolar tDCS increased functional connectivity between the mPFC and right sFG and mFG and decreased negative functional connectivity between the mPFC and the right anterior insula and basal ganglia. Analyses of intrinsic functional networks revealed that frontopolar tDCS reduced functional connectivity between the SN and DMN. Frontopolar tDCS also accelerated therapeutic learning during an individualized ERP-like extinction exercise in patients with OCD. These findings suggest that targeted brain stimulation may modulate functional connectivity and can potentiate therapeutic learning during exposure-based CBT. If so, these effects might be harnessed to accelerate or enhance the clinical response to psychotherapy.

The simplest explanation of the behavioral findings is that frontopolar anodal tDCS directly modulates neuronal excitability in the frontal pole and mPFC,^51, 88^ thereby promoting intracellular processes in the mPFC that support LTP-like plasticity recruited during extinction learning.^89-92^ It is also possible that frontopolar stimulation remotely modulated excitability or LTP-like plasticity of other regions implicated in fear extinction, outside the region of direct stimulation. This latter idea is compelling given that functional connectivity changed between the frontal pole and several other areas implicated in fear extinction learning – though not commonly included in the canonical fear circuit – following administration of tDCS. These include the right dlPFC, the right anterior insula, anterior putamen, ventral caudate, and ventral pallidum.^93^ For example, the dlPFC, which has been shown to be functionally connected to the vmPFC,^94^ is important for the explicit regulation of emotions during fear extinction training.^95^ These interpretations are speculative as these processes were not directly measured, and fMRI connectivity analyses and the behavioral study were performed in distinct groups of participants.

Many of the brain regions identified in our seed-based fMRI connectivity analysis are contained within three canonical brain networks: the DMN, the SN, and the right ECN; **Figure 2** and **Supplemental Figure 3**). Analysis of ICA components suggested that connectivity between the SN and DMN was most affected by tDCS. The observed effects of frontopolar tDCS on therapeutic learning may be mediated by modulation of inter-network relationships. SN activation – particularly in the right anterior insula – has been found to be triggered by salient cues, to inhibit the DMN, and to activate the ECN.^96-98^ In the present study, the frontal pole seed region (which is within the DMN) and the right anterior insula cluster (which is within the SN) were anticorrelated at baseline but moved toward zero functional connectivity following tDCS. Similarly, anticorrelations between the SN and all three DMN components were significantly reduced following tDCS. This reduced anticorrelation may have raised the threshold at which SN activation could inhibit or “switch off” the DMN. Since DMN activity has been implicated in safety learning, reduced DMN inhibition during the processing of emotionally arousing and salient stimuli (e.g., a contaminated object) may promote the processing and learning of safety signals during extinction.^37^ Again, this is speculative, since we did not measure DMN activity during our therapeutic extinction learning task.

Extinction learning is typically considered a form of implicit learning, but explicit emotion regulation and learning processes are also important.^95^ These explicit processes are commonly associated with ECN activation.^95, 99^ Increased functional connectivity between the frontal pole and dlPFC – an anchor of the ECN – following administration of tDCS may have increased the probability that these regions are co-activated, thus promoting both DMN-dependent implicit safety learning and ECN-dependent emotion regulation or explicit learning during therapeutic exposure. This interpretation is not supported by analyses of ICA networks, which showed no significant changes in connectivity between the right frontoparietal network and the SN or DMN.

Unexpectedly, we also observed significant reductions in functional connectivity between the mPFC and multiple areas in the subcortical reward circuitry, particularly in the striatum, following administration of frontopolar tDCS (**Figure 2A**; **Supplemental Figure 2**). This includes areas associated with fear extinction learning and the detection of safety signals and related approach behaviors.^100,101^ The two striatal networks identified in ICA analyses showed no significant changes in connectivity following frontopolar tDCS. However, the SN, which showed reduced connectivity following tDCS, included striatal areas that overlapped with the right anterior insula/putamen cluster identified in seed-based analyses. Elevated striatal activity is seen in individuals with OCD, both at rest and upon symptom provocation, and is moderated by treatment.^102, 103^ In our patient sample, tDCS may have modulated activity in cortico-striatal-thalamic loops and associated obsessive thinking or compulsive impulses, which might make the ERP Challenge less distressing. Future research should examine this idea in the context of fMRI-based OCD symptom provocation paradigms.^104, 105^

Although promising, these findings have several important limitations. First, sample sizes were relatively small for both studies; replication with larger samples is necessary. Second, imaging and behavioral data were collected in distinct samples. This was necessary because our behavioral extinction task was designed to recapitulate procedures used during CBT for OCD and was not compatible with MRI, but it weakens linkage of the two studies. Third, sample characteristics were not uniform across the two studies. The neural effects of frontopolar tDCS in community and OCD samples may differ. Ideally, future research should attempt to integrate stimulation, imaging, and therapeutic exposure into one protocol to allow investigators to examine if changes in functional connectivity mediate the effects of frontopolar tDCS on therapeutic learning. Lastly, study one utilized a within-subject design and lacked a control group. While within-subject functional connectivity is largely stable over time, particularly for fMRI protocols that implement reliability safeguards used in the present study (e.g., eyes open, awake, short re-test intervals),^106^ a well-powered, double-blind, sham-controlled fMRI study is needed to make stronger inferences about the causal effects of frontopolar tDCS on functional connectivity. Study 2, conversely, utilized a double-blind, sham-controlled design. The OCD subject and the experimenter administering the ERP Challenge were both blind to the experimental condition, but the experimenter administering tDCS was not always blind.

Predicted difficulty of exposures during exposure planning were nearly identical across the two groups, as were SUDS ratings immediately before and after tDCS administration and during the first several minutes of the first exposure trial. Nonetheless, replication efforts should ensure blinding of all individuals involved in data collection efforts.

These limitations aside, the present findings are both novel and promising. To the authors’ knowledge, these are the first studies to examine *both* behavioral and neurobiological target engagement of tDCS in the context of therapeutic exposure. The present findings have clear implications for the treatment of anxiety and related disorders, including OCD. If replicated with larger samples and extended with samples of other anxious populations and over multiple sessions of tDCS and exposure, this strategy may lead to a novel means to accelerate recovery, reduce dropout, and improve response rates in the psychotherapeutic treatment of OCD, and perhaps of other disorders characterized by anxious pathology.

## Data Availability

Raw study data are available upon request from the corresponding author.

## Acknowledgements

Dr. Adams and the reported studies were supported by the National Institute of Mental Health (NIMH; K23MH111977, T32MH062994, and L30MH111037). Study one was also supported, in part, by the Clinical Neurosciences Division of the National Center for PTSD and the Department of Veteran Affairs. Brain stimulation equipment was loaned by Starstim® to support study pilots and was purchased using funds from the Detre Foundation (R13306). Study two was supported, in part, by an International Obsessive-Compulsive Disorder Foundation (IOCDF) Young Investigator Award and an American Psychiatric Association (APA) Psychiatric Fellowship Award (R12965). These studies were also supported by the State of Connecticut through its support of the Ribicoff Research Facilities at the Connecticut Mental Health Center. Dr. Anticevic is supported by the NIMH (R01MH108590). Dr. Abdallah is supported by the Beth K. and Stuart Yudofsky Chair in the Neuropsychiatry of Military Post Traumatic Stress Syndrome. Portions of these data were presented at annual conferences for the American College of Neuropsychopharmacology (ACNP) and Anxiety and Depression Association of America (ADAA). Dr. Pittenger is supported by the Taylor Family Foundation and the NIMH (R01MH116038 and K24MH121571). The views in this article are those of the authors, not of the State of Connecticut or of other funders. We would like to thank Eileen Billingslea, MA, and Suzanne Wasylink, APRN, for their assistance with IRB compliance, subject recruitment and screening, and budget management.

