## Supplemental Materials for "Transcranial direct current stimulation (tDCS) targeting the medial prefrontal cortex (mPFC) modulates functional connectivity and enhances inhibitory safety learning in obsessive-compulsive disorder (OCD)"

*MRI Sequences*

After localization, high-resolution anatomical images were collected: T1W MPRAGE (TR=2400ms, TE=2.01ms, TA=7 min., FOV=256mm, voxel=0.8 mm^3^) and T2 SPACE (TR=3200ms, TE=565ms, TA=6 min., FOV=256mm, voxel=0.8 mm^3^). An HCP-style multiband fMRI sequence was used for rs-fMRI acquisition, parameters were as follows (TR = 700 ms, TE = 31 ms, FOV = 210 mm, voxel = 2.2 mm^3^).

*fMRI Processing*

Image preprocessing followed standard steps and was completed using AFNI software as previously reported.^1^ Anatomical (T1) images were skull stripped (SSWarper), segmented into CSF, WM, and GM (3dSeg), and then warped into MNI space (3dNwarpApply). EPI images underwent despiking, deobliquing, motion correction using rigid body alignment, alignment to participant’s normalized anatomical images, detrending, bandpass filtering (.01 and .1 hz), spatial smoothing using an 8 mm FWHM Gaussian filter (AFNIs 3dBlurToFWHM that estimates the amount of smoothing to add to each dataset to result in the desired level of final smoothing), and rescaling into percent signal change. Images were normalized using the MNI 152 template brain. Following recent recommendations, we corrected for head motion related signal artifacts by using motion regressors derived from Volterra expansion, consisting of [R R2 Rt-1 R2t-1], where R refers to each of the 6 motion parameters, and separate regressors for mean signal in the CSF and WM.^2, 3^ This step was implemented directly after motion correction and normalization of the EPI images in the image preprocessing stream. One participant was removed from second-level analyses due to excessive artifacts and problems with alignment.

*Measures*

*Dimensional Obsessive Compulsive Scale* (DOCS) is a 20-item self-report measure that assesses severity of four symptom dimensions of OCD [5-items each: contamination, responsibility, unacceptable thoughts, and symmetry] over the past month.^4^ *Generalized Anxiety Disorder – 7* (GAD-7) is a 7-item self-report measure used to assess levels of trait anxiety over the last 2 weeks.^5^ *Patient Health Questionnaire* (PHQ-9) is a 9-item self-report measure that assesses severity of depression over the last 2 weeks.^6^ *Subjective Units of Distress* SUDS ratings were anchored to patient-specific examples of no distress (0), moderate distress (50), and extreme distress (100).The *tDCS* *Adverse Effects Questionnaire* (tDCS-AEQ) assesses for the presence of ten common tDCS side effects.^7^ Presence of side effects are rated on a scale from zero (absent) to three (severe). If a participant endorsed the presence of a side effect, they were then asked to rate the degree to which they believed the side effect was related to tDCS using a scale from zero (“none”) to four (“definitely”). The final question on the tDCS-AEQ was used to assess the degree to which each participant believed they received active (real) or sham (placebo) stimulation using a bi-polar scale anchored at zero (“not sure”) and ranging from -2 (“definitely sham”) to +2 (“definitely active”).

*Exposure and Response Prevention (ERP) Challenge*

On Day 1 of the ERP Challenge, patients were first provided standardized ERP psychoeducation.^8^ They then worked with an experimenter to develop a brief hierarchy of candidate exposure exercises. This was done to limit inflation or deflation of estimates of exposure difficulty; initial estimates of difficulty for proposed exposures were often very high or low but were adjusted accordingly once considered relative to other exposure options. When indicated, the experimenter would demonstrate exposure exercises to clarify techniques. The experimenter and patient then selected a single exposure exercise, which was used for the remainder of the ERP Challenge. See **Supplemental Table 2** for example exposure exercises.

Five 10-minute trials of the selected exposure exercise were completed, with approximately one-minute breaks between trials. SUDS were assessed every minute before, during, and after each exposure trial. Response prevention compliance was assessed for each trial and was scored on a 0 (no compulsions) to 5 (extreme compulsions) scale based on experimenter observations and patient report. At the end of Day 1, the patient was encouraged to continue response prevention to the best of their ability until their next appointment. Day 2 of the ERP Challenge commenced 18-36 hours later in the same room with the same experimenter. The session began with a brief interview to establish response prevention adherence between sessions. The patient then completed the same exposure exercises as on Day 1.

*Seed-Based Analyses of fMRI Data*

An anatomical mask of the frontal pole was extracted from the Desai DKD maximum probability maps^9^, which overlapped significantly with the areas most effected by the anode (compare **Figures 1a** and **2a**). We then extracted the average BOLD time course within the frontal pole using 3dmaskave for each participant and each of their four runs. This timecourse was then regressed separately onto each remaining grey matter voxel’s timecourse using 3dDeconvolve and 3dREMLfit. Fisher’s *r*-to-*z* transformation (3dcalc) was used to improve normality and resulting *z-*values were used to calculate each participant’s whole-brain functional connectivity seedmap representing the degree to which each voxel was correlated with the frontal pole seed over the timecourse.

All reported effects appeared on the right side of the brain. To test whether this is truly a lateralized effect or is simply an artifact of thresholding, we conducted exploratory analyses with relaxed voxelwise correction (cluster size of 30.6 and uncorrected *p* < .01). The main effect of tDCS was significant at the *p* < .01 level for sixteen clusters (see **Supplemental Figure 1**). In addition to the previously listed clusters in the right hemisphere, functional connectivity increased between the frontal pole and the left superior and middle frontal gyri, and negative connectivity decreased between the frontal pole and left basal ganglia. This suggests that most effects of tDCS are likely bilateral, though statistically stronger on the right. Functional connectivity between the frontal pole and other nodes in the DMN and SN also changed following administration of tDCS. These included, but were not limited to, clusters in right parietal lobe and rostral anterior cingulate cortex (rACC), which increased in connectivity, and clusters in the dorsal anterior cingulate cortex (dACC) and supplementary motor area (SMA), which decreased in connectivity.

To better map the anatomy of the subcortical structures that reduced their negative functional connectivity after tDCS, a mask of the two significant clusters in the basal ganglia was extracted and mapped onto a mask of reward circuitry. Negative functional connectivity decreased throughout the anterior striatum, including the putamen and the head and body of the caudate, the globus pallidus (interior, exterior, and ventral), and the nucleus accumbens. Functional connectivity also decreased in the right medial hypothalamus and the extended amygdala. See **Supplemental Figure 2**

A mask of the cortical regions that increased in functional connectivity following administration of tDCS was extracted and mapped onto masks of the DMN, SN, and executive control network (ECN).^10^ The frontopolar seed mapped exclusively onto the DMN. The sFG and mFG clusters both mapped primarily within the ECN, with some medial overlap with DMN. The anterior insula cluster mapped primarily within the SN, with some overlap with the ECN. See **Supplemental Figure 3**.

*Independent Component Analysis*

Independent component analysis (ICA) was implemented with the infomax algorithm within the Group Independent Component Analyses for fMRI Toolbox (GIFT).^11^ GIFT parameters included: Group ICA (GICA) using ICASSO with options set to include random initializations and 50 ICA decomposition iterations to establish solution reliability, and back reconstruction of spatial component maps using GICA3; removal of mean voxel values in pre-processing; and principle components analyses (PCA) with singular value decomposition (SVD) accomplished with two reduction steps (90 and 35), single precision, and selective eigen solver. The resulting 35 components were scaled to z-scores.

*Day 2 ERP Challenge Learning*

A final LME model was used to examine potential between-group differences in within- and between-trial learning during Day 2 of the ERP Challenge; LME parameters were identical to those described above for Day 1 (see **Supplemental Table 3**)**.** Importantly, no tDCS was administered on Day 2, so any group differences or interactions derive from the effects tDCS delivered on Day 1. OCD patients in the active tDCS group reported lower SUDS ratings than those in the sham tDCS group during the first minute of exposure on Day 2 (group *β* = -12.69, 95%C.I. = -24.77 to 0.61). Significant main effects of mins (*β* = -1.01, 95%C.I. = -1.72 to -0.30) and trials (*β* = -3.41, 95%C.I. = -5.53 to -1.29) reflected continued decline in SUDS ratings throughout Day 2. Despite neither group receiving tDCS on Day 2, those in the active tDCS group continued to report accelerated within-trial learning relative to those in the sham tDCS group. (group*mins *β* = -1.11, 95%C.I. = -2.12 to -0.11). However, the rate of between-trial learning was similar across the two experimental groups (see **Supplemental Figure 4**; trial *β* = 0.26, 95%C.I. = -2.74 to 3.26).

**Supplemental Table 1.** Background data for Study 1 volunteers (*n* = 18) and Study 2 OCD patients (*n* = 24).

|  | **Study 1** | **Study 2** | |
| --- | --- | --- | --- |
|  | **Active-tDCS** | **Sham-tDCS** (*n* = 12) | **Active-tDCS** (*n* = 12) |
| Age | *M* = 36.44, SE = 3.06 | *M* = 32.42, *SE* = 3.67 | *M* = 34.17, *SE* = 4.32 |
| Gender | 44% female | 83% female | 66% female |
| Race | 55% Caucasian | 75% Caucasian | 75% Caucasian |
| Medication | 0% Psychotropic | 38% Psychotropic | 32% Psychotropic |
| DOCS | *M* = 2.72, *SE* = 0.68 | *M* = 32.23, *SE* = 4.98 | *M* = 27.23, *SE* = 3.95 |
| GAD-7 | *M* = 1.44, *SE* = 0.45 | *M* = 15.42, *SE* = 17.72 | *M* = 17.5, *SE* = 1.84 |
| PHQ-9 | *M* = 0.90, *SE* = 0.25 | *M* = 7.33, *SE* = 2.04 | *M* = 9.17, *SE* = 2.15 |

***Note****.* All between-group t-tests or *Χ^2^* tests for Study 2 were non-significant (all *ps* > .10). DOCS = Dimensional Obsessive-Compulsive Scale, GAD-7 = Generalized Anxiety Disorder Scale, and PHQ-9 = Patient Health Questionnaire.

**Supplementary Table 2.** Example exposure and response prevention (ERP) exercises completed during the ERP Challenge.

| **Fear/Obsession** | **Exposure** | **Response Prevention** | **Predicted SUDS** | **Actual SUDS** |
| --- | --- | --- | --- | --- |
| Will be responsible for starting a fire in home | Use blow dryer, turn off, then leave on top of papers | No visual checking or reassurance seeking | 50 | 50 |
| Will curse ex-girlfriend with negative thoughts | Write her name repeatedly while thinking “bad” thoughts | No self-assurance or praying | 45 | 50 |
| Fear of catching flu-like illness and vomiting | Touch cloth to doorknob then hold cloth against cheek | No wiping or washing | 55 | 45 |
| Need for exactness and symmetry with personal belongings | Pull items (e.g., license) out of wallet, place back imperfectly, then stare | No ordering or arranging and mental neutralizing | 40 | 60 |

**Note.** Minor details changed or removed for privacy. SUDS = Subjective Units of Distress (0-100).

**Supplementary Table 3.** Linear mixed effects modeling of within- (mins) and between-trial (trials) learning during Day 2 of exposure with response prevention (ERP) challenge among OCD patients who received Active-tDCS (*n* = 12) or Sham-tDCS (*n* = 12) approximately 24 hours prior. Significant main effects of mins and trials suggest that SUDS ratings continued to decline throughout Day 2 of the ERP Challenge. A significant main effect of group suggests that OCD patients in the Active-tDCS group reported lower SUDS ratings than those in the Sham-tDCS group during the first trial of the first exposure on Day 2. The significant mins*group interaction suggests that, despite neither group receiving tDCS on Day 2, those in the Active-tDCS group continued to report accelerated within-trial SUDS reductions relative to those in the Sham-tDCS group (see **Supplemental Figure 4**).

|  |  | **95% C.I.** | |
| --- | --- | --- | --- |
| **Fixed Factors** | **Beta (SE)** | **Lower** | **Upper** |
| Intercept | 47.80 (4.14) | 39.26 | 56.34 |
| Mins | -1.01 (0.34)*** | -1.72 | -0.30 |
| Trials | -3.41 (1.03)*** | -5.53 | -1.29 |
| Group | -12.69 (5.86)* | -24.77 | -0.66 |
| Mins*Trials | 0.10 (0.06)*** | -.01 | 0.21 |
| Group*Mins | -1.11 (0.49)* | -2.12 | -0.11 |
| Group*Trials | 0.26 (1.46) | -2.74 | 3.26 |
| Group*Mins*Trials | 0.17 (0.08)* | 0.01 | 0.32 |

**Note:** * = *p* < .05; *** = *p* < .001. SUDS = Subjective Units of Distress

**Supplemental Figure 1.** Main effects of tDCS (pre-to-post tDCS) with cluster correction (30.6 voxels) and relaxed thresholding (uncorrected *p* < .01) suggested that, following administration of tDCS, functional connectivity between the frontal pole (red): increased with the rostral anterior cingulate cortex and the bilateral middle and superior frontal gyri, and; decreased with the bilateral basal ganglia, right anterior insula, and dorsal anterior cingulate cortex.

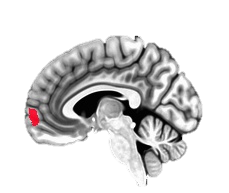

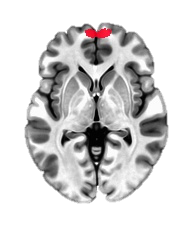

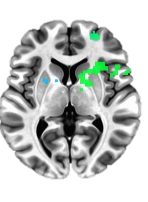

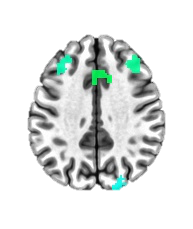

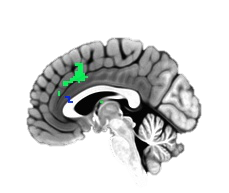
**Supplemental Figure 2.** Seed-based functional connectivity analyses with stringent cluster correction (*p* < .001, cluster sizes > 30.6 voxels) suggested that tDCS decreased functional connectivity between the frontal pole and reinforcement circuitry, particularly the right anterior basal ganglia, including the head and body of the caudate, putamen, globus pallidus (ventral, interior and posterior), nucleus accumbens, hypothalamus, and extended amygdala.

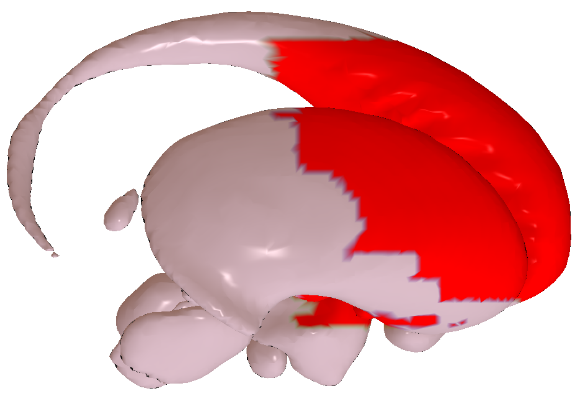

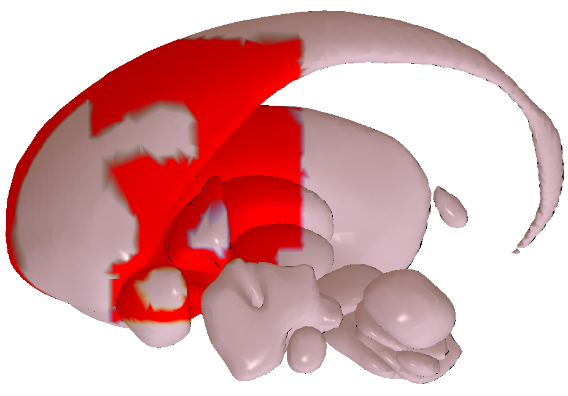

**Supplemental Figure 3.** The frontal pole seed and the cortical clusters in which tDCS significantly altered functional connectivity (red) mapped onto default mode network (DMN; yellow), salience network (SN; green-blue), and executive control network (ECN; blue). Masks extracted from the CAREN atlas ^10^ suggest that the entire frontopolar seed and much of the mFG were within the DMN (orange), the majority of the anterior insula cluster and the anterior portion of the sFG cluster are in the SN (red-green), and the remainder of the sFG cluster and nearly all of the mFG cluster are in the ECN (purple).

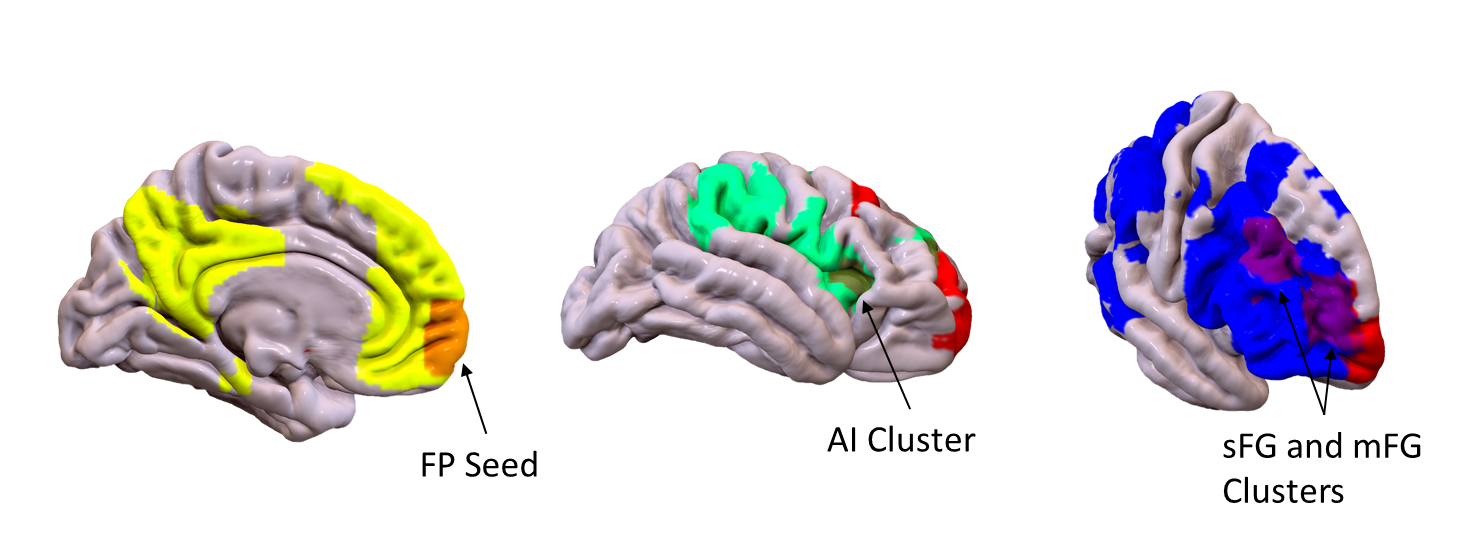

**Supplemental Figure 4**. **A.** SUDS ratings did not significantly differ between the two experimental groups from Study 2 before or after tDCS (*p*s > .10). **B.** Participants who received Sham-tDCS reported being similarly convinced that they received Active (real) tDCS as those who received Active-tDCS (*p*=.92), suggesting that Sham tDCS was a credible placebo.

**A.**

SUDS

|  | Sham-tDCS | Active-tDCS |
| --- | --- | --- |
| Pre-tDCS | 35.13 | 59.38 |
| Post-tDCS | 36.25 | 36.88 |

**B.**

| Definitely  Placebo (Sham)  tDCS | Not Sure | Definitely  Real (Active)  tDCS |
| --- | --- | --- |

|  | Column1 |
| --- | --- |
| Sham | 0.5 |
| Active | 0.57 |

**Supplementary Figure 5.** Linear mixed effects modeling suggested that OCD patients who received Active-tDCS on Day 1 of the experiment reported significantly (*p* < .05) lower SUDS ratings at the beginning of Day 2 (min. 1 of trial 1) compared to those who received Sham-tDCS on Day 1. Despite no administration of tDCS on Day 2 of the ERP Challenge, OCD patients who received Active-tDCS 24 hours earlier continued to evidence significantly (*p* ≤ .01) faster within-trial learning (solid lines) relative to those who received Sham-tDCS the day prior. Unlike Day 1 of the ERP Challenge, however, there were no group differences in between-trial learning (dashed lines).

Average SUDS Ratings

| Day 2 ExposureTrials | Sham | Active |
| --- | --- | --- |
| 1 | 50.25 | 38.9167 |
|  | 48 | 35.6667 |
|  | 44.0833 | 33.5 |
|  | 43.3333 | 29.8333 |
|  | 42.3333 | 27.6667 |
|  | 42.6667 | 25.3333 |
|  | 42.8333 | 23.8333 |
|  | 41.3333 | 23 |
|  | 40.8333 | 21.25 |
|  | 40.1667 | 20.6667 |
| 2 | 44.1667 | 32.3333 |
|  | 40.75 | 27.75 |
|  | 40.6667 | 25 |
|  | 40.5 | 19.6667 |
|  | 39.75 | 18.0833 |
|  | 38.3333 | 18.1667 |
|  | 38.3333 | 16.4167 |
|  | 38.6667 | 15.9167 |
|  | 37.9167 | 16.1667 |
|  | 37.0833 | 15.25 |
| 3 | 41.25 | 27.5833 |
|  | 40.0833 | 25.75 |
|  | 38.9167 | 23.9167 |
|  | 38.0833 | 21.5 |
|  | 37.75 | 18 |
|  | 36.25 | 18.3333 |
|  | 35.3333 | 16.75 |
|  | 33 | 15.8333 |
|  | 32.4167 | 16.5 |
|  | 32.4167 | 15.8333 |
| 4 | 36.5455 | 24.8333 |
|  | 35.3636 | 25.3333 |
|  | 35.2727 | 22.0833 |
|  | 35.4545 | 20.5 |
|  | 34.5455 | 19.6667 |
|  | 35.0909 | 18.5833 |
|  | 35.0909 | 16.9167 |
|  | 33.0833 | 16.6667 |
|  | 33.25 | 16.1667 |
|  | 32.4167 | 16.5 |
| 5 | 33.75 | 24.3333 |
|  | 33.5 | 22.25 |
|  | 33.4167 | 21.6667 |
|  | 32.5833 | 19.0833 |
|  | 33.1667 | 18.75 |
|  | 31.3333 | 17.75 |
|  | 30.6667 | 16.9167 |
|  | 29.4167 | 15.9167 |
|  | 29.1667 | 15.4167 |
|  | 29.5833 | 14.75 |

***Note****.* SUDS = Subjective Units of Distress (0-100).
